# A stronger association of depression with rheumatoid arthritis in presence of obesity and hypertriglyceridemia

**DOI:** 10.1101/2023.01.01.23284106

**Authors:** Grayden Shand, Daniel T. Fuller, Leon Lufkin, Carly Lovelett, Nabendu Pal, Sumona Mondal, Shantanu Sur

## Abstract

**Background:** Rheumatoid arthritis (RA) is an autoimmune disorder characterized by chronic and systemic inflammation. Recent research underscores the role of chronic inflammation in multiple common RA comorbidities such as depression, obesity, and cardiovascular diseases (CVDs), suggesting a potential overlap of the pathogenic mechanisms for RA. However, it is not well understood how the coexistence of these comorbid conditions impacts the risk of RA and whether any such association relates to the inflammatory status of the body.

**Methods:** We used data from the 2007-2010 United States National Health and Nutrition Examination Survey (NHANES) database and compared RA prevalence between subsamples with the presence of any two conditions among depression, obesity, and hypertriglyceridemia (HTG). Each subsample was further divided into three categories based on the serum level of the inflammatory marker C-reactive protein (CRP) and analyzed for statistically significant differences using three-way χ^2^ tests of independence.

**Results:** The study was conducted on 4,136 patients who fulfilled the inclusion criteria (representing 163,540,241 individuals after adjustment for sampling weights). Rates of depression, obesity, and HTG were found to be significantly higher (*P* < 0.001) among the subjects with RA compared with the control population with no arthritis. The presence of depression along with obesity or HTG showed a noticeably higher RA prevalence but such an association was not observed for the combination of obesity and HTG. The synergistic effect of HTG with depression was found to be most prominent at a medium CRP level (1-3 mg/L), while for obesity, the effect was observed across all CRP levels examined. These findings were further confirmed by the three-way χ^2^ test for independence.

**Conclusions:** The presence of obesity or HTG in subjects suffering from depression might pose an increased risk of RA. Inflammatory mechanisms potentially play an important underlying role as suggested by the strong dependency of the association to CRP level. Identification of synergistic associations between RA risk conditions could provide useful information to predict the development and progress of RA.

## Introduction

Rheumatoid Arthritis (RA) is a chronic progressive autoimmune disease affecting an estimated 0.5-1% of adults in North America.^1,2^ It primarily affects the synovial joints and is characterized by stiffness, major joint pain, and irreversible joint deformity causing serious disability if left untreated. The prevalence of RA varies across age, gender, and ethnicity with a number of genetic, environmental, and behavioral factors known to increase disease risk.^1,3–6^ RA often associates with a variety of comorbid conditions including depression, obesity, cardiovascular diseases (CVDs), and diabetes,^7–10^ suggesting an overlap of RA pathogenic mechanisms with these chronic conditions. The existence of a shared or overlapping pathway of pathogenesis would not only increase the prevalence of comorbidities among RA patients but could potentially increase the risk of RA when one or more comorbidities are present in a patient. Since chronic, systemic inflammation with dysregulated immunity is a hallmark of RA and underlies most of its clinical manifestations, we speculate that comorbidities with a known inflammatory link would be highly pertinent in this context.^11^ While many different comorbidities among RA subjects have been studied in detail, less information is available on whether the presence of multiple comorbidities poses a risk for the development or progress of RA.

Depression is identified as a major RA comorbidity with a prevalence of 13-42% in RA patients and is associated with a poorer outcome with increased pain, disability, and risk of mortality.^12–14^ Analyzing longitudinal data from the National Health Insurance Research Database of Taiwan, Lu et al. reported that the incidence of depression among RA patients was 15.69 per 1,000 person-years (PYs) compared with 8.95 per 1,000 PYs in the control population.^15^ Interestingly, this study also found the incidence of RA to be significantly higher in the population with depression than in the control population, suggesting a bidirectional relationship between RA and depression.^15^ Similar to depression, obesity is reported to have a higher prevalence among RA patients, and thus, given the adverse impact of obesity on multiple chronic diseases, one major focus of research is to understand how obesity influences the course of RA.^16,17^ Studies have linked obesity to worsened disease outcomes in RA and also found that obese patients had lower odds of achieving and sustaining remission when compared with non-obese patients.^18,19^ Although the role of obesity on RA development has not been established fully, the results from several longitudinal studies indicate obesity increases the risk of RA.^17^ Other conditions such as hypertriglyceridemia (HTG), a condition when the serum triglyceride concentration exceeds a certain level (150 mg/dL), are also important from the context of their interactions with other RA-associated conditions.^20^ Specifically, HTG is associated with increased atherosclerosis formation,^21^ and is a well-known risk factor for many CVDs.^22^ Risk of several CVDs including coronary artery disease and peripheral vascular disease as well as major CVD events such as myocardial infarction and stroke are substantially increased in RA, resulting in higher CVD-related death.^23–25^ While HTG has not been shown to be an important risk factor for RA, it is associated with increased severity of symptoms, potentially related to its relationship to CVDs.

One major hallmark of RA is chronic inflammation associated with a dysregulation of the innate and adaptive immune systems.^26^ Stimulation of inflammatory pathways has also been reported for depression, obesity, and HTG. For example, excess fat tissue in obese individuals is associated with high levels of inflammatory cytokines, including tumor necrosis factor alpha (TNF-α), interleukin 1 (IL-1) 1, interleukin 6 (IL-6), and C-reactive protein (CRP), all of which are known mediators of RA.^27,28^ Similarly, higher levels of TNF-α, IL-1, IL-6, and CRP are reported in subjects with increased depression.^29^ HTG is also associated with increased inflammation (manifested by increased levels of CRP, IL-6, and other inflammatory markers),^30^ which is partially attributed to a reduced anti-inflammatory capacity of serum high-density lipoproteins resulting from a higher-than-average triglyceride content.^31^ The higher level of inflammatory markers in these conditions and RA suggest inflammation plays an important role in the development of these diseases. Moreover, the frequent occurrence of these conditions in subjects with RA is suggestive of a common molecular underpinning potentially mediated through inflammatory mechanism.^26^

The objective of this work is to investigate any potential relationship between depression, obesity, and HTG – each of which independently relates to chronic inflammation – in their association with RA. Using publicly available data from National Health and Nutrition Examination Survey (NHANES), we explored whether the coexistence of any two of these conditions synergistically enhances the association with RA. Considering their common connection with an inflammatory mechanism, we also studied if such association is related to the general inflammatory status of the subject.

## Materials & Methods

### Study Population

The CDC-operated online portal NHANES offers detailed health data, obtained through interviews and physical examinations, on samples drawn from the US population. It employs a stratified, multistage, nationally representative cross-sectional survey design in order to provide countrywide inference. The survey protocol and data collection methods of NHANES data used in this study were approved by the National Center for Health Statistics Ethics Review Board (protocol #2005-06). The types of data collected by NHANES include demographic variables, socioeconomic condition (SEC), questionnaires, and bio-specimen examination. The participants are represented by a unique number in each dataset to protect their identity.

### Data Collection

Analyses were conducted on data from the 2007-2008 and the 2009-2010 NHANES datasets to examine the relationship of inflammation (measured by CRP) with the three primary factors of interest: depression, obesity, and triglyceride. These years were chosen because of their detailed information on CRP, which is not present in later years of NHANES data. Response rates for participation in both interviews and physical examinations were close for each two-year cycle and ranged from 75% to 80%. Subjects younger than 18 years were excluded from the analysis to prevent inclusion of juvenile RA and subjects with ages greater than 79 years were excluded as NHANES does not differentiate ages 80 years or above.

### Variables and Measurements

#### RA and Comorbid Conditions

##### RA

RA was defined based on answers to the following questions: (1) “Has a doctor or other health professional ever told that you had arthritis?” If the response to this question is yes, the second question was (2) “What type of arthritis?” Subjects with the response of “Rheumatoid Arthritis” were considered to have RA.

##### Depression

A nine-item depression screening instrument built on Patient Health Questionnaire (PHQ-9) was used to assess depression. PHQ-9 scores can range from 0 to 27 and we considered subjects with a score of ≥ 10 to suffer from depression, which includes the categories of moderate-severe, and severe depression.^32^

##### Hypertension

Three consecutive individual blood pressure measurements 30 seconds apart were obtained by certified examiners using a sphygmomanometer after the participants had been seated and had rested for at least five minutes. A fourth measurement was taken if any of the previous three were missing or performed erroneously. The means of these recorded values were used to represent the participants’ systolic and diastolic blood pressures. Systolic and diastolic hypertension were defined when the measured systolic and diastolic blood pressure were ≥130 mm Hg and ≥80 mm Hg, respectively.

##### Hypertriglyceridemia (HTG)

Triglyceride measurements were obtained with the Roche/Hitachi Modular P Chemistry Analyzer method. Serum triglyceride concentration ≥ 150 mg/dL was considered as HTG in accordance with standards set by the Adult Treatment Panel III of the National Cholesterol Education Program.^20^

##### Obesity

Height was measured using a wall-mounted stadiometer, and weight was measured using a Toledo digital scale. BMI was calculated from standing height and body weight measurements. Participants with BMI ≥ 30 kg/m^2^ were considered obese.

##### Diabetes

Participants were recorded as diabetic if they self-reported the condition or had hemoglobin A1c ≥ 6.5%. Self-reported diabetes was determined from the participant answering “yes” to at least one of a set of questions regarding physician diagnosis and insulin regulation.

##### Stroke

Stroke was defined as a self-reported history of stroke.

#### Other Variables

##### CRP

CRP measurements in serum samples were obtained using latex-enhanced nephelometry. Measured CRP levels were further divided into three risk categories of low (< 1 mg/L), medium (1-3 mg/L), and high (> than 3 mg/L) based on the guideline provided by the Centers for Disease Control and Prevention (CDC) and the American Heart Association (AHA).^33^

##### Demographics

Age, sex, and median family incomes were obtained from the NHANES database.

### Statistical Analysis

Each data point of the NHANES data was adjusted by implementing appropriate sampling weight according to the National Center for Health Statistics analytical guidelines to account for the potential bias created by the sampling procedure.^34^ We used weights for triglycerides, the most discriminatory dataset used in our study, and then adjusted them to account for the inclusion of both the 2007-2008 and the 2009-2010 datasets. We have presented here the analyses both with and without the implementation of sample weights.

All *P*-values presented here are from two-sided tests. *P*-values less than 0.05 were considered statistically significant. Inter-group difference in abundance was evaluated using a Z-proportion test for dichotomous indicators and a two-way chi-square (χ^2^) test was used for non-dichotomous indicators. Three-way χ^2^ tests were used to test for significant differences in group proportions of subpopulations for the weighted data. Residual analysis was utilized to identify the degree of contribution to the test statistic from each categorical combination.

Data preprocessing and statistical analyses were performed using R software version 3.2.5 and all plots were generated using the ggplot2 package.

## Results

NHANES study included 10,149 participants in 2007-2008 and 10,537 participants in 2009-2010. Out of these 20,686 participants from 2007-2010, the inclusion criteria were satisfied by 4,136 subjects (Figure 1), of which 305 were positive for RA. A comparison of demographics and major comorbidities between the RA and non-RA study population (unweighted *n* = 4,136; weighted *N* = 163,540,241) is shown in Table 1. Age and gender distributions were found to be significantly different between these two groups. 50.2% of the RA population belonged to the age group of more than 60 years and 40% belonged to the age group of 41-60 years, while for the non-RA population these numbers were 18% and 35%, respectively. Consistent with the literature, females were more commonly affected by RA with 61% of RA subjects being female compared with 49% female population in the non-RA group. The percentage of the population belonging to the lower income group (annual income < $25K) was higher among RA subjects (42.6%) than non-RA subjects (30.8%).

**Figure 1.**
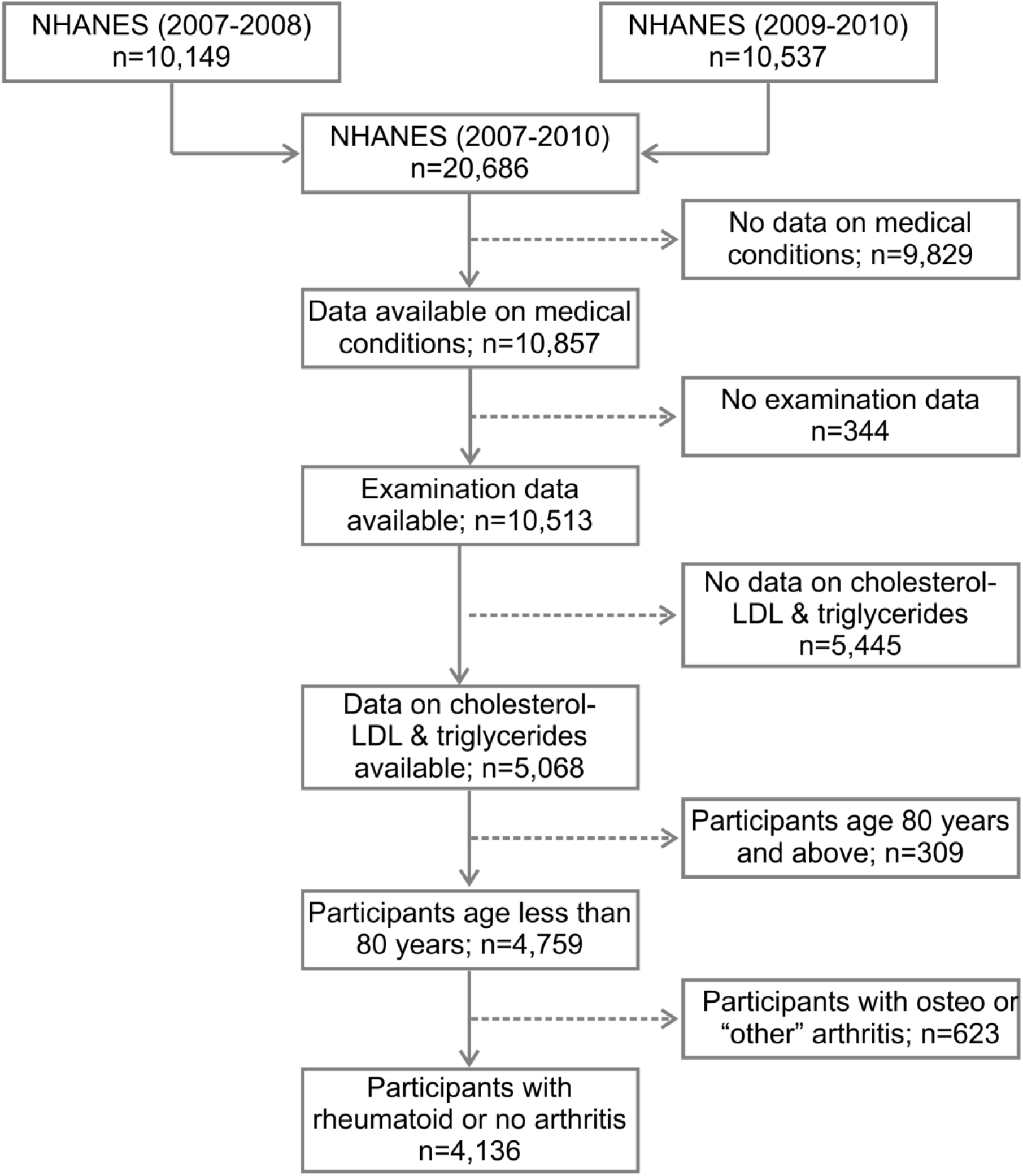
Flow diagram of the selection of study population from NHANES data (2007-2010). NHANES, National Health and Nutrition Examination Survey; LDL, low-density lipoproteins.

**Table 1.** Comparison of study populations. Moderate to severe depression is considered a comorbidity for RA in the table. Systolic and diastolic blood pressures are used to assess hypertension. The symbol * indicates a χ2 test was performed while the symbol † denotes a two-sample proportion Z-test.

| Variables | Unweighted<br>*n* = 4,136 |  | Weighted<br>*N* = 163,540,241 |  | P |
| --- | --- | --- | --- | --- | --- |
|  | No Arthritis<br>*n* = 3,831 (%) | RA<br>*n* = 305 (%) | No Arthritis<br>*N* = 155,157,729 (%) | RA<br>*N* = 8,380,512 (%) |  |
| **Age** |  |  |  |  | <0.0001* |
| ≤ 40 | 1,781 (46.5) | 30 (9.8) | 78,833,815 (50.8) | 1,148,070 (13.7) |  |
| 41-60 | 1,358 (35.5) | 122 (40) | 58,410,172 (37.6) | 3,879,670 (46.3) |  |
| > 60 | 692 (18) | 153 (50.2) | 17,915,741 (11.5) | 3,352,772 (40.0) |  |
| **Gender** |  |  |  |  | <0.0001* |
| Male | 1,957 (51.1) | 120 (39.3) | 79,796,903 (51.4) | 3,371,900 (40.2) |  |
| Female | 1,874 (48.9) | 185 (60.7) | 75,362,826 (48.6) | 5,008,613 (59.8) |  |
| **Income** |  |  |  |  | <0.0001* |
| < \$25,000 | 1,180 (30.8) | 130 (42.6) | 35,684,140 (23.0) | 3,126,096 (37.3) | |
| \$25,000-75,000 | 1,471 (38.4) | 109 (35.7) | 60,118,646 (38.8) | 3,187,188 (38.0) | |
| > \$75,000 | 819 (21.4) | 41 (13.4) | 50,108,916 (32.3) | 1,474,647 (17.6) | |
| NA | 361 (9.4) | 25 (8.3) | 9,248,027 (6.0) | 592,582 (7.1) |  |
| **Comorbidities** |  |  |  |  |  |
| Depression | 246 (6.4) | 61 (20.0) | 8,299,320 (5.4) | 1,576,573 (18.8) | <0.0001† |
| Hypertriglyceridemia | 493 (12.9) | 63 (20.7) | 18,444,010 (11.9) | 1,610,680 (19.2) | <0.0001† |
| Obesity | 1,292 (33.7) | 157 (51.5) | 448,086,197 (31.0) | 4,529,270 (54.1) | <0.0001† |
| Stroke | 64 (1.7) | 29 (9.5) | 1,679,696.2 (1.1) | 691,128.8 (8.3) | <0.0001† |
| Diabetes | 319 (8.3) | 84 (27.5) | 8,787,123 (5.7) | 2,003,661 (23.9) | <0.0001† |
| Hypertension (sys.) | 755 (20.7) | 124 (40.7) | 22,579,296 (14.6) | 2,936,593 (35.0) | <0.0001† |
| Hypertension (dias.) | 593 (15.5) | 42 (13.8) | 21,935,832 (14.1) | 1,438,295 (17.2) | <0.0001† |

Individuals with RA were found to have a stronger association with common comorbidities including hypertension, stroke, hypertriglyceridemia, depression, diabetes, and obesity (Table 1; *P* < 0.0001 for all). The distributions of the three RA comorbidity indicators considered here (PHQ-9 score, BMI, and triglyceride level) were further visualized among RA and non-RA populations using violin plots (Figure 2). For all three measurements, we observed a distinct difference in the plot shape between the two groups with a higher probability density at higher values for the RA population. This observation indicates that RA is associated with a distributional shift to higher values of PHQ-9 score, BMI, and triglyceride level.

**Figure 2.**
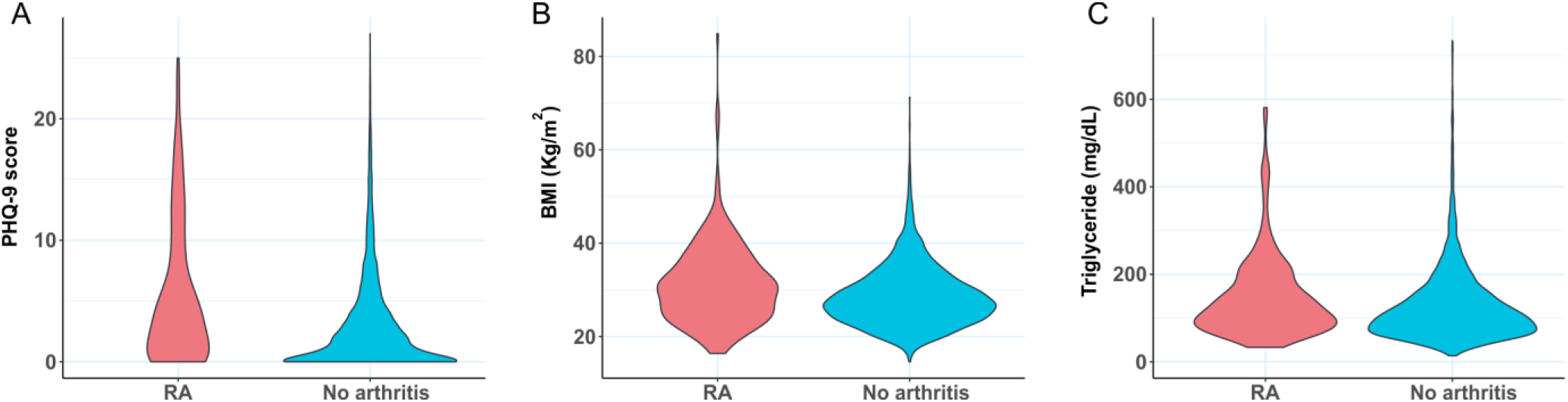
Violin plots comparing the distribution of (A) depression (PHQ-9 score), (B) BMI, and (C) triglyceride level between RA subjects and control with no arthritis. BMI, body mass index; PHQ-9, Patient Health Questionnaire (9 questions related to patient health).

Tissue inflammation being a hallmark of RA, we compared serum levels of inflammatory marker CRP between RA and non-RA populations. The cumulative distribution plot of CRP showed a clear right shift for the RA population when compared with the plot for the control population, suggesting higher inflammation (Figure 3A). For example, according to this plot, 70.2% of the control population has CRP levels below 3.0 mg/L value, while only 47.7% of the RA population falls below this concentration. The reported association of chronic low-grade inflammation with depression, obesity, and HTG also motivated us to explore any connection of CRP with PHQ-9 score, BMI, and triglyceride level. ^35,36^ We divided each of these three variables into two categories based on medically relevant threshold values, separating subjects with depression (PHQ-9 score ≥ 10), obesity (BMI ≥ 30), and HTG (triglyceride ≥ 150 mg/dL) from subjects without these conditions (Figure 3B-D). Cumulative distribution plots of CRP revealed higher values in populations with depression, obesity, and HTG with the difference being most prominent for obesity. The analysis of CRP level thus supports the reported connection of inflammation with RA as well as the comorbid conditions considered in this study.^30,31^

**Figure 3.**
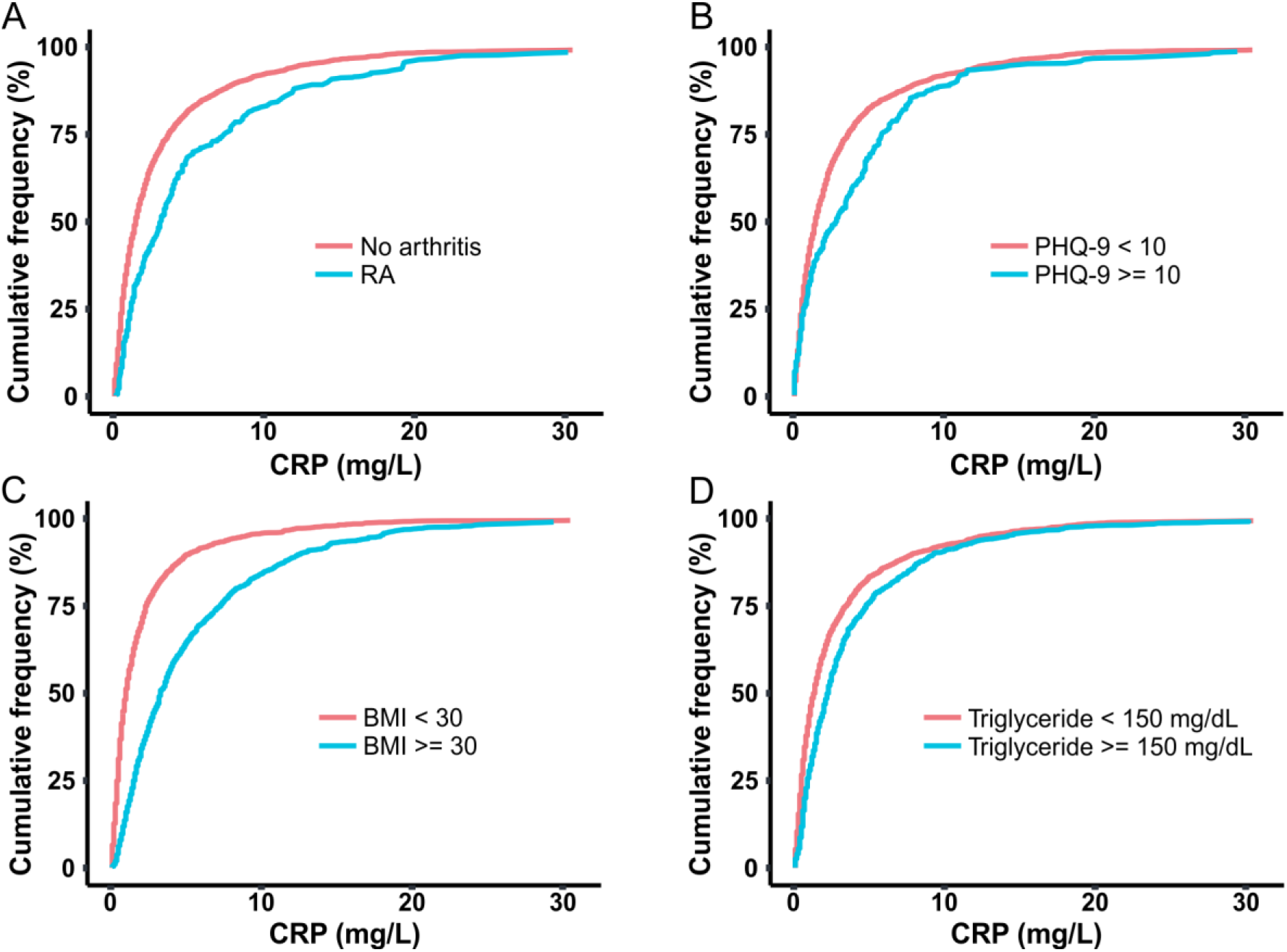
Cumulative distribution plots comparing CRP level among populations of (A) RA and no arthritis, (B) moderate-severe and no-mild depression (PHQ-9 score cut off at 10), (C) high and low BMI value (cut off at 30 kg/m^2^), (C) high and low triglyceride concentration (cut off at 150 mg/dL), and (D). CRP, C-reactive protein; BMI, body mass index; PHQ-9, Patient Health Questionnaire (9 questions related to patient health).

Next, we explored how the presence of these comorbidities, alone or in combination, influences the association with RA. Since we observed subjects with both RA as well as in subjects with depression, obesity and HTG have higher CRP values we were interested to understand whether any such association is impacted by the inflammatory state. To investigate this question, we divided the sample population into three categories based on CRP levels of low (< 1 mg/L), medium (1-3 mg/L), and high (> 3 mg/L). This categorization divided the total sample population into three groups of comparable size. We observed the percentage of RA subjects progressively increase from low to high CRP group (Figure 4), further confirming the connection of inflammation to RA.

**Figure 4.**
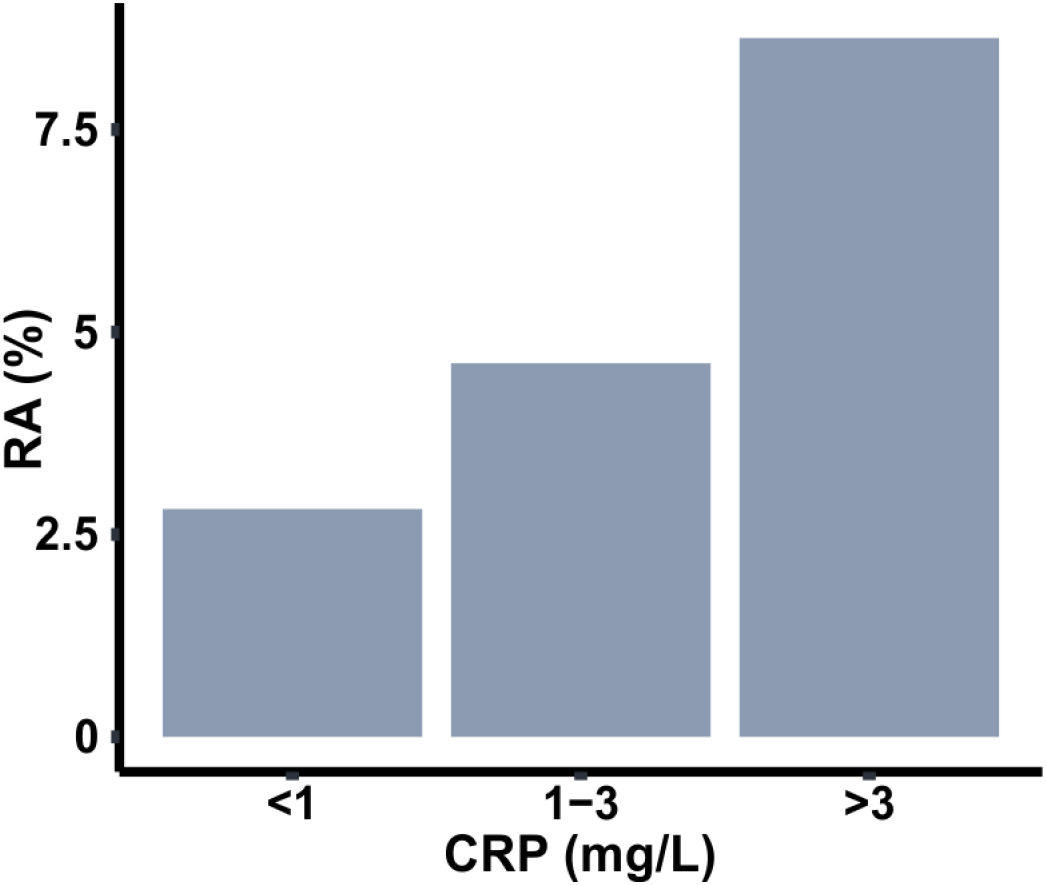
Prevalence of RA in among populations with three different ranges of C-reactive protein (CRP).

Since the subjects with depression, obesity, and HTG showed higher CRP values in cumulative distribution plots, we studied their prevalence across the CRP groups. Specifically, we were interested to know whether the presence of one of these conditions could influence the coexistence of another condition. To explore this, we created 2×2 tile plots in each of the CRP ranges under consideration, taking any two conditions at once (Figure 5). Colormap representation of the sample population in these pair-wise tile plots revealed that the majority of the population in the low CRP group consisted of the absence from either condition. The proportion of subjects with obesity increased substantially at medium and high CRP levels; HTG showed a similar but weaker trend, but such a change was not conspicuous for depression. Next, we checked the pattern of coexistence between any two conditions, where a higher propensity for coexistence will be reflected by a relatively higher proportion of subjects in the upper right quadrant of a 2×2 tile plot. On examination of the tile plots, no preferential association could be observed across the CRP groups.

**Figure 5.**
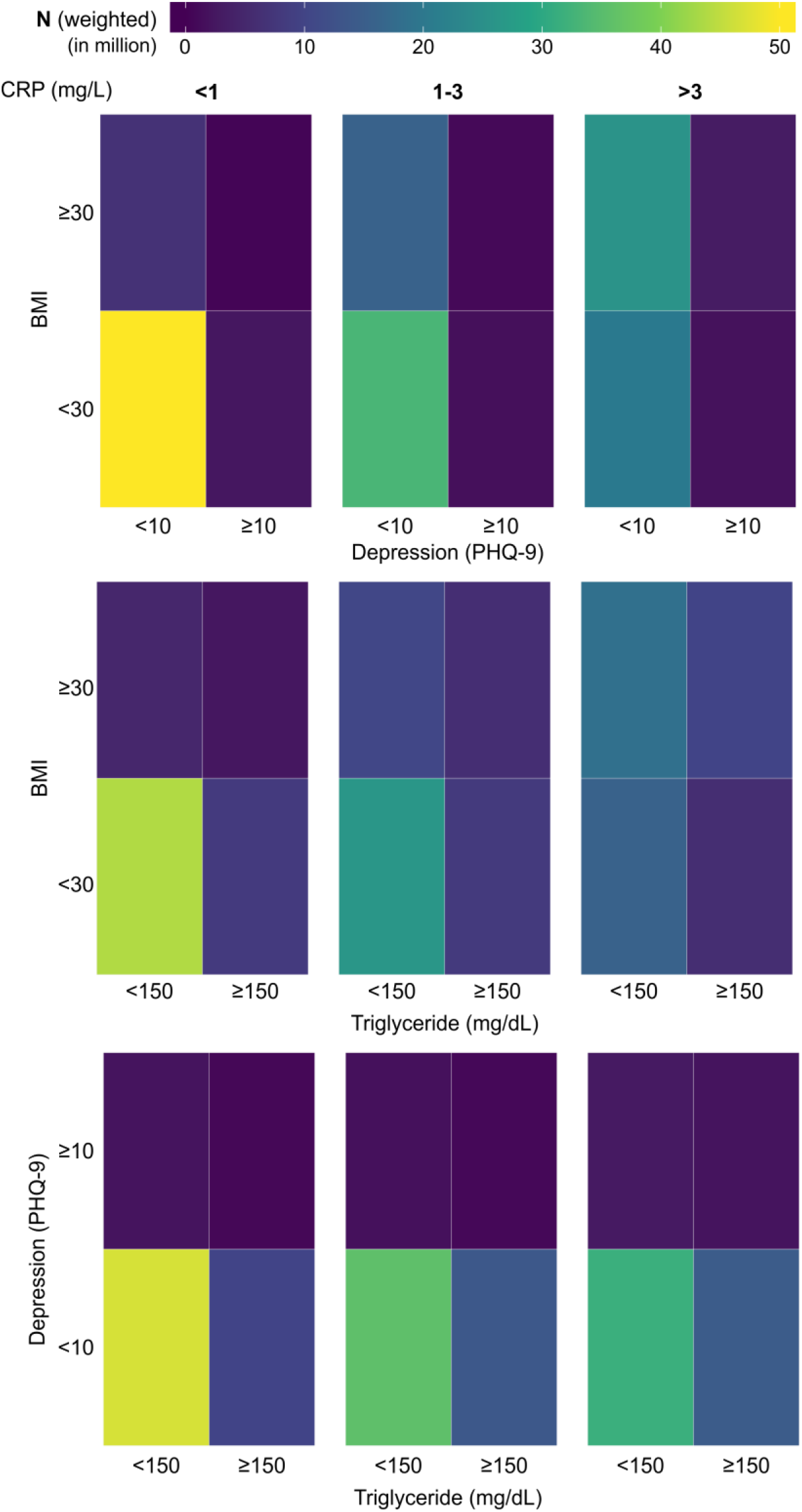
Tile plots showing the coexistence of depression, high BMI, and high triglyceride in the population, considering any two variables at a time. The population is further subdivided into three categories based on CRP level (<1 mg/L, 1-3 mg/L, and >3 mg/L). CRP, C-reactive protein; BMI, body mass index; PHQ-9, Patient Health Questionnaire (9 questions related to patient health).

We took a similar approach to examine whether the coexistence of any two conditions among depression, obesity, and HTG could influence the association with RA. Similar to Figure 5, 2×2 tile plots were created but the colors here indicated the percentage of subjects with RA in each category (Figure 6). The plots revealed distinct differences in RA prevalence depending upon the presence of comorbidities and CRP levels. Depression or obesity was found to be associated with a noticeable increase in RA prevalence, but such a relationship was not observed for HTG. Furthermore, for obesity, the effect was more prominent for the subjects in the low and medium CRP group, while for depression, a stronger effect was observed in the medium or high CRP group. Additionally, we observed a substantial increase in RA prevalence among subjects who have depression along with obesity or HTG. This effect was observed across all CRP ranges for obesity with a slightly stronger effect at the medium level of CRP (1-3 mg/L); for HTG, the effect was prominent only at the medium CRP level. For example, when depression and obesity were considered at the medium CRP range, the prevalence of RA in subjects with depression and obesity were 19.5% and 5.5%, respectively; however, RA prevalence increased to 27.2% for the population with both depression and obesity. Similarly, a high RA prevalence of 34.9% was observed among subjects with depression and HTG at the medium CRP level, while the prevalence of RA among subjects with depression and HTG alone were 16.3% and 5.6%, respectively. Interestingly, at a high CRP level (>3 mg/L), the coexistence of HTG among the subjects with depression had a lower prevalence of RA (12.9%) than subjects with depression but not HTG (22.9%).

**Figure 6.**
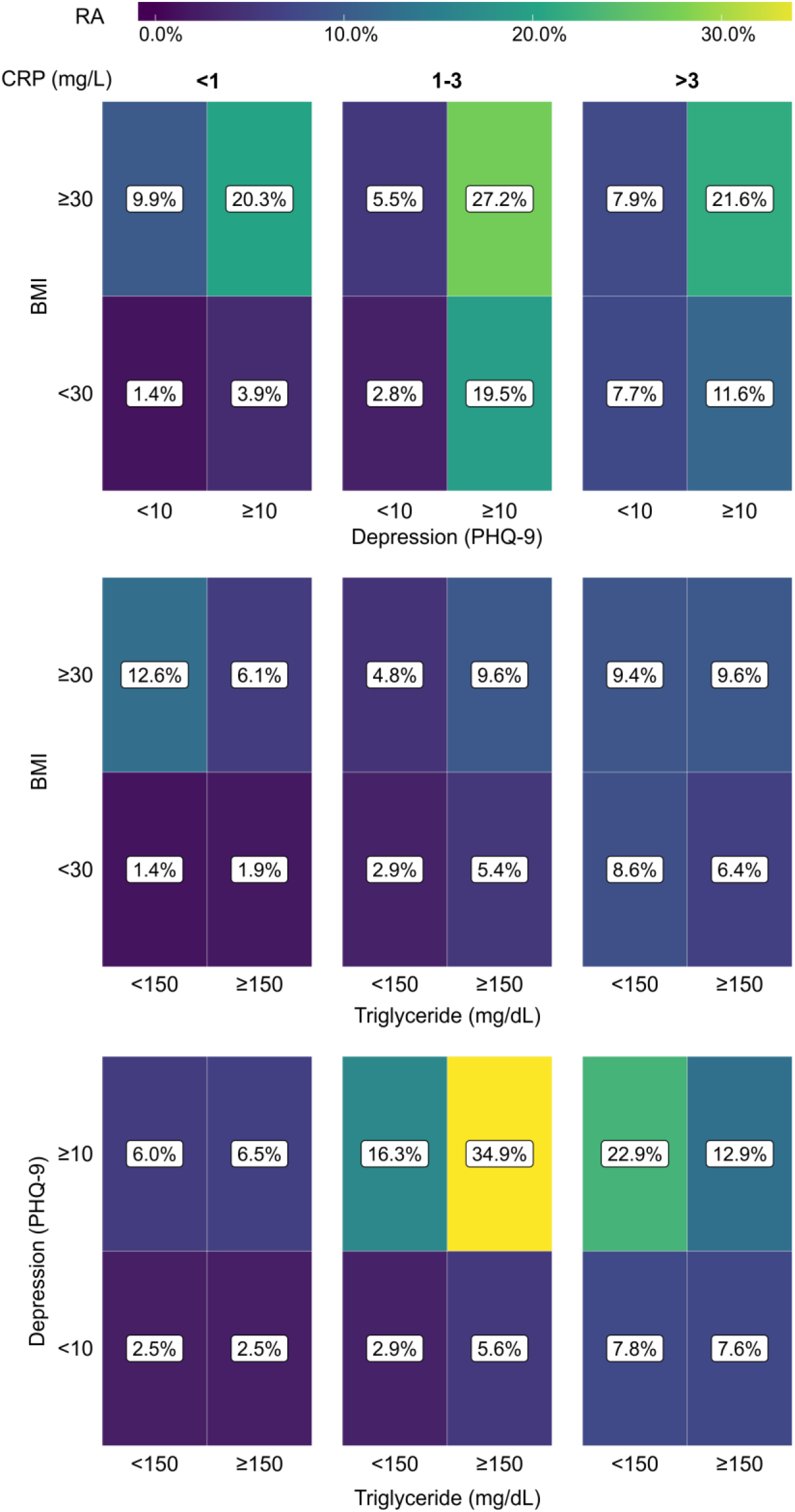
Tile plots showing the prevalence of RA (%) in the population with coexistent presence of depression, high BMI, and high triglyceride considering any two comorbidities at a time. The population is further subdivided into three categories based on CRP level (<1 mg/L, 1-3 mg/L, and >3 mg/L). CRP, C-reactive protein; BMI, body mass index; PHQ-9, Patient Health Questionnaire (9 questions related to patient health).

To make a quantitative interpretation of the findings in Figure 6, we conducted three-way χ^2^ tests of independence (Table 2). We find the proportion of subjects with RA to be significantly different for all situations, except when depression and HTG were considered at the low level of CRP. The standardized residuals were then used to understand the contribution of each categorical combination to the χ^2^ test result. Corroborating the observation from Figure 6, high standardized residual values were found for RA positivity when depression coexisted with obesity at all CRP levels or with HTG in the medium level of CRP. For example, among the subjects with depression and HTG in the medium level of CRP, the standardized residuals for RA positive and negative were 6.35 and -0.76, respectively; in contrast, the standardized residual values for RA positive and negative were 0.97 and -0.34 when depression with the absence of HTG was considered in the same CRP range. Furthermore, when the presence of the same pair of conditions was considered at the high CRP level, the standardized residual values for RA positive and negative were 1.75 and 1.28, respectively. Thus, the χ^2^ test results confirm the synergistic relationship of depression with obesity and HTG for the association with high RA prevalence, and potential contribution of the inflammatory states of the subjects to underlie such association.

**Table 2.**
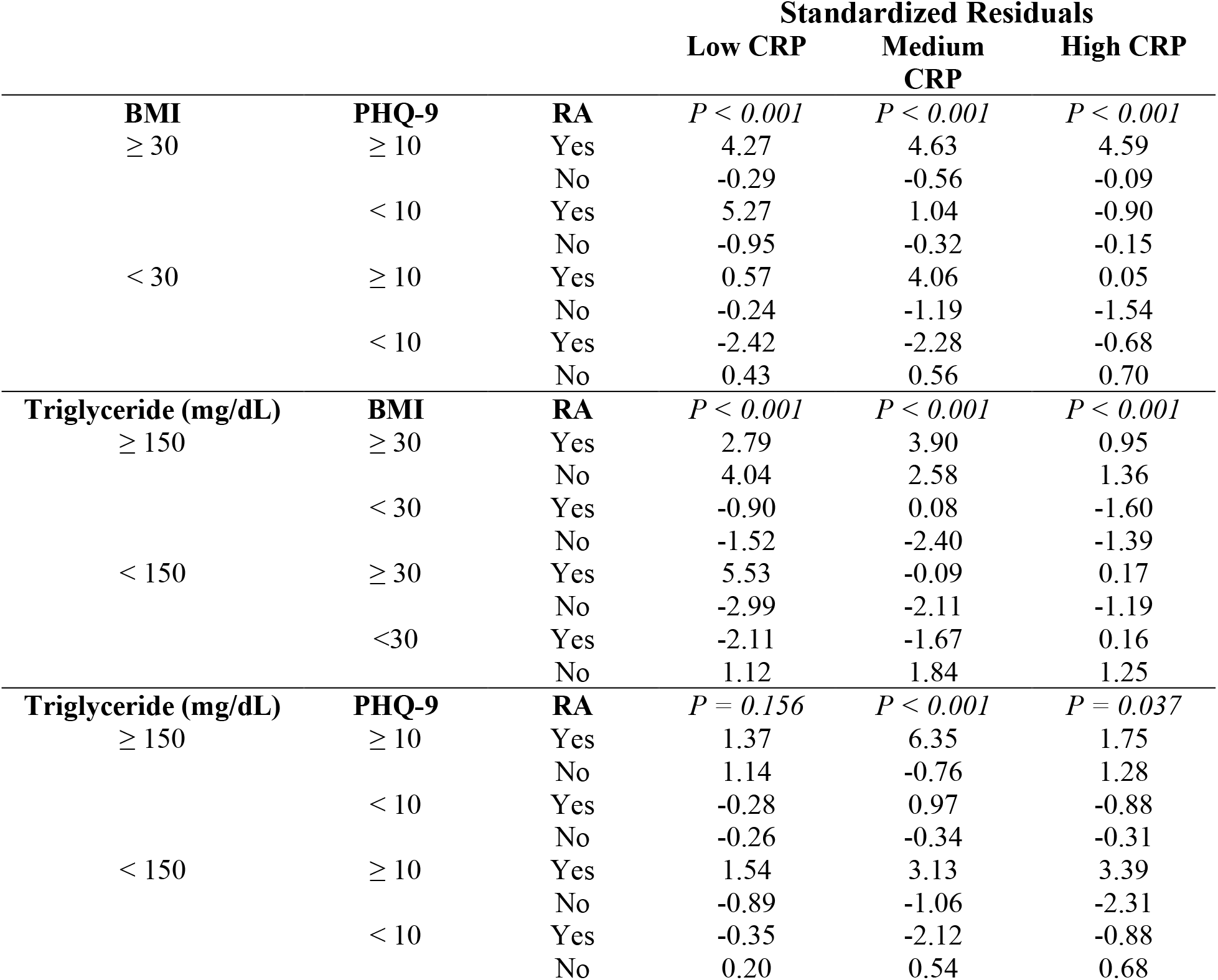
Standardized residuals from χ2 tests for the association between depression, obesity, and hypertriglyceridemia in RA measured at three different plasma CRP levels (low, < 1 mg/L; medium, 1-3 mg/L, and high, > 3 mg/L).

## Discussion

In this study, we compared the association of RA with three common chronic conditions depression, obesity, and HTG with a specific focus on any changes in their association due to the coexistence of another condition. We observed that the prevalence of RA among subjects with depression was strongly increased in the presence of obesity or HTG. Our analysis also showed that the degree of synergistic association is strongly dependent on the inflammatory state of the body with the effect being maximum at the medium level of CRP (1-3 mg/L).

The relationship between depression and RA has been extensively studied since it is one major comorbid condition for RA and significantly impacts the disease outcome, including mortality.^12–14^ Recent studies suggest a bidirectional association between these two disease conditions, implying that subjects suffering from depression are also more likely to develop RA.^15,37^ Our study indicates that the proportion of RA among subjects with depression is considerably higher when the subjects also had obesity or HTG. Interestingly, even though RA prevalence was higher among subjects with obesity, the coexistence of HTG did not increase the prevalence. Thus, the observed synergistic association is specific to depression among the conditions studied here. Due to the cross-sectional nature of the data, we can’t conclude whether the presence of obesity or HTG poses an increased risk of developing RA among the subjects with depression. Also, the size of our study population did not allow us to study if the RA prevalence is further impacted when both obesity and HTG are present in subjects with depression as the number of subjects with all three conditions was too low to perform any meaningful analysis. Regardless, our findings imply the need for additional attention when evaluating depression with obesity or HTG and warrant a longitudinal study to evaluate the risk of RA in these subjects.

One key finding from our analysis is that the observed association is dependent on CRP level, a marker of inflammation. CRP is widely used in clinical settings to evaluate RA disease activity,^38^ and we have also found RA prevalence to increase with the CRP level. The reported connection of inflammation with depression, obesity, and HTG is also corroborated in our analysis where CRP levels were found to be higher in these conditions compared with the control populations.^27–30^ Interestingly, the CRP levels at which obesity and HTG influenced the association of depression with RA were found to be distinct. While the influence of BMI was observed across all CRP levels (low, medium, and high), the effect of HTG was prominent only at the medium CRP level. Furthermore, HTG was found to decrease the prevalence of RA among subjects with obesity at low CRP level and subjects with depression at high CRP level. Overall, these results suggest a potentially important role of immunity in determining the interactions between depression, obesity, and HTG on their association with RA; however, the nature of such interactions is not well understood from the current results based on CRP. Measurement of CRP can be very useful to quickly assess the systemic inflammatory state, but it is highly nonspecific, and its serum level can be elevated by a broad array of conditions that induces inflammation, including infections and tissue damage. Inflammation involves a complex interplay of large numbers of inflammatory mediators and specific pathways are known to be predominant in the pathogenesis and manifestation of certain disease conditions. Therefore, a deeper exploration of the inflammatory pathways is necessary to better understand the associations observed here.

In recent years, there is an emerging interest in the prediction of RA from a set of known risk factors. Such interest is primarily motivated by the detection of the disease at the “pre-RA” period when the subject is yet to clinically develop RA symptoms, but the presence of RA-related autoantibodies can be detected in the blood.^39,40^ Diagnosis in the pre-RA period, although challenging, can be extremely advantageous as intervention started at an earlier stage can considerably slow down the disease progress and bring favorable outcomes.^39^ Models constructed toward RA prediction considered a wide range of potential risk factors under consideration, including genetic, socio-demographic, and environmental factors, comorbidities, and results of clinical and laboratory examination.^41–45^ However, interactions between these factors were not included in most of these models – except when a specific interaction is already known or suspected^41,44^ – as that would otherwise exponentially increase the computational cost.^45^ In this context, our observation of synergistic interactions of depression with obesity and HTG could provide important insights to incorporate these specific interactions in a prediction model to improve prediction accuracy.

Although our analysis provides important insights into how the coexistence of certain chronic conditions can influence their association with RA, there are a few limitations in this study. First, the cross-sectional nature of the data only allows the inference on the association between variables rather than concluding on their causal relationship. Given the subjects of depression are at higher risk of RA, our findings highlight the need for a longitudinal study investigating the impact of the coexistence of depression with BMI or obesity on RA incidence. Second, the self-reported diagnosis of RA used in this study is prone to overestimate RA prevalence due to the increased chance of false-positive diagnosis, mostly coming from erroneous inclusion of other forms of arthritis. While well-established diagnostic criteria of RA such as one provided by the American College of Rheumatology (ACR) and European League Against Rheumatism (EULAR)^46^ should be used for clinical decision-making, self-reported RA is considered to have an acceptable level of accuracy for epidemiological studies.^47^ Since NHANES provides a large nationwide dataset with a very well-structured sampling design, we believe that even with these limitations, it provides suitable data to conduct preliminary analysis. Finally, CRP is a good marker for the initial screening of the inflammatory state but has a very limited value in probing the potential crosstalk between inflammatory signaling pathways. Therefore, to understand the molecular underpinnings of the synergistic interactions observed here, measurement of a broader panel of inflammatory markers (e.g., TNF-α, IL-1, IL-6) should be incorporated in future studies.

## Data Availability

Publicly available datasets were analyzed in this study. This data can be downloaded from: https://wwwn.cdc.gov/nchs/nhanes/

## Acknowledgments

Daniel T. Fuller acknowledges the support from the Lawrence ‘57 and Antoinette Delaney Ignite Research Fellowship.

## Notes

### Competing Interest Statement

The authors have declared no competing interest.

### Funding Statement

This study did not receive any funding

### Author Declarations

Publicly available human data were analyzed in this study. The data were openly available before the initiation of the study. This data can be downloaded from: https://wwwn.cdc.gov/nchs/nhanes/

